# Outcomes following acute poor-grade aneurysmal subarachnoid bleed - Is early definitive treatment better than delayed management?

**DOI:** 10.1101/2020.06.10.20127316

**Authors:** Adam Gittins, Nick Talbott, Ahmed A Gilani, Greg Packer, Richard Browne, Randeep Mullhi, Zaheed Khan, T Whitehouse, Antonio Belli, Rajnikant L Mehta, Fang Gao-Smith, Tonny Veenith

## Abstract

**Background/Objective:** Patients with poor-grade subarachnoid bleed (pSAH, World Federation of Neurosurgeons grades 4-5) often improve their neurocognitive function months after their ictus. However, it is essential to explore the timing of intervention and its impact on long-term outcome. We compared the long-term outcomes between immediate management within 24 hours (IM) and delayed management after 24 hours (DM) in patients following pSAH.

**Methods:** This was a retrospective population-based study, including patients with pSAH who received definitive management between 1^st^ January 2011 and 31^st^ December 2016 in a large tertiary neurocritical care unit. The primary outcome was adjusted odds ratio (OR) of favourable outcome (Glasgow Outcome Scale (GOS) 4-5) for survivors at 12 months following discharge, as measured by the Glasgow Outcome Scale (GOS). The secondary outcomes included adjusted OR of a favourable outcome at discharge, three months and six months following discharge and survival rate at 28 days, three months, six months and 12 months following haemorrhage.

**Results:** 111 patients were included in this study: 53 (48%) received immediate management (IM) and 58 (41%) received delayed management (DM). The mean time delay from referral to intervention was 14.9±5.8 hours in IM patients, compared to 79.6±106.1 hours in DM patients. At 12 months following discharge, the adjusted OR for favourable outcome in IM versus DM patients was 0.96 (CI 0.17, 5.39; p=0.961). At hospital discharge, three months and six months, the adjusted OR for favourable outcome was 3.85 (CI 1.38, 10.73; p=0.010), 1.04 (CI 0.22, 5.00; p=0.956) and 0.98 (CI 0.21, 4.58; p=0.982), respectively. There were no differences in survival rate between the groups at 28 days, three months, six months and 12 months (71.7% in IM group vs 82.8% in DM group at 12 months, p=0.163).

**Conclusions:** IM and DM after pSAH are associated with similar morbidity and mortality at 12 months. Therefore, delaying intervention in poor-grade patients may be a reasonable approach, especially if time is needed to plan the procedure or stabilise the patient adequately.

## INTRODUCTION

The optimal time to secure an aneurysm after a poor-grade aneurysmal subarachnoid bleed (pSAH) is debatable ^1^. pSAH is defined as World Federation of Neurosurgeons (WFNS) grades 4-5 after the subarachnoid bleed (SAH). The International Subarachnoid Aneurysm Trial (ISAT) has established the safety and efficiency of endovascular aneurysm coiling as a treatment in patients with a good-grade (WFNS grades 1-3) subarachnoid bleed.^2,3^ To date, the majority of data on the outcomes after an acute SAH have been on patients with good clinical grades (WFNS grades 1-3) at presentation. Data are required to optimally manage the 18-24% of patients presenting with pSAH.^4^ The Treatment of Poor-Grade Subarachnoid Haemorrhage Trial 2 (TOPSAT2) study, which attempted to redress this balance by enrolling patients with pSAH, has stopped recruiting due to slow recruitment (personal communication).^5^

The timing of pSAH management depends on the surgical risk, availability of skilled personnel, risk of re-bleed, and uncertainty regarding neurocognitive disability after a pSAH. These factors have led to variable practices within the UK, ranging from early management within hours to delayed management after a pSAH.^6-8^ Investigations such as digital subtraction angiography, lack of ‘round the clock’ interventional neurovascular and radiology services, and management of reversible causes of low Glasgow Coma Score (GCS) such as hydrocephalus often delay treatment after a pSAH.

Advances in surgical and endovascular techniques^9^ with provision of high quality of critical care^4^ have substantially lowered the risk of morbidity and mortality in pSAH. A systematic review comparing outcomes after pSAH based on the timing of intervention demonstrated that early and intermediate surgery (within days 0-3 and days 4-7, respectively) offered better outcomes than late surgery.^10^

Recently, small retrospective and prospective centre cohort studies have shown encouraging results after early aggressive treatment of pSAH.^11-14^ These data have challenged the widespread belief that the prognosis of pSAH is ‘too poor’ to intervene.^11,15-17^ Our cohort study aims to compare the outcomes of pSAH after early definitive treatment (within 24 hours) to delayed treatment (after 24 hours).

## METHODS

### Setting

All regional referrals with aneurysmal SAH to our tertiary referral neurosciences centre between January 1^st^ 2011 and December 31^st^ 2016 were considered for inclusion in this study. This research used anonymised linked databases and was classed as a service evaluation (CARMS-14561) with a waiver of ethical approval according to the national guidelines (http://www.hra-decisiontools.org.uk/research/docs/definingresearchtable_oct2017-1.pdf) as supported by our institution.

### Patient Identification

All regional referrals to our tertiary referral centre were logged into a National On-Call Referral Service (NORSe) database with details on patient demographics, clinical presentation, diagnosis, and investigation results. These data, along with the linked information from electronic patient records and general practice records, were used for this study. All patients referred to a tertiary neurovascular centre between 1^st^ January 2011 and 31^st^ December 2016 in the West Midlands with a poor-grade aneurysmal SAH (WFNS grade 4-5, at the time of referral) were included in this study. Patients were excluded if they had good-grade SAH (WFNS grade 1-3) on referral, non-aneurysmal haemorrhage (e.g. traumatic or infective cause for SAH) or had suffered cardiac arrest after the initial bleed^18^ and if they did not receive definitive treatment to secure the aneurysm.

Baseline characteristics such as patient demographics, Glasgow Coma Scale (GCS) score, Charlson Comorbidity Index (CCI) score, and nature and timing of intervention received were retrieved from the neurosciences electronic patient records. Primary and secondary outcome data were collected from electronic patient records.

### Outcomes

The objective was to compare immediate management within 24 hours of referral (IM) to delayed management after 24 hours (DM). The primary outcome was adjusted odds ratio (OR) of favourable outcome, defined as Glasgow Outcome Scale^19^ (GOS) 4-5, at 12 months, calculated during the patient follow-up. The secondary outcomes were adjusted OR of favourable outcome at discharge, three months and six months following discharge and survival rate at 28 days, three months, six months and 12 months following haemorrhage.

### Statistical Analysis

Statistical analyses were conducted using Statview (Version 5, 1998, SAS Institute Inc., Cary, North Carolina, USA). Baseline characteristics of groups were presented as mean (standard deviation), median (25%, 75% quartile) and percentages. Comparison between groups on continuous data was analysed using the unpaired *t*-test. When the assumption of normality was violated, non-parametric techniques such as the Mann-Whitney test were used. The Chi-squared test was used for comparison between percentages and frequencies. We used binary logistic regression to estimate the effects of IM and DM on the dichotomous GOS measures at hospital discharge, three months, six months and 12 months respectively. Furthermore, we adjusted management effect (IM vs DM) for baseline covariates in the logistic regression (age, gender, CCI, GCS and intervention received). The results were considered statistically significant when *p*-value <0.05.

## RESULTS

### Patient Characteristics

A total of 1525 NORSe referrals for suspected aneurysmal SAH were made in the years 2011-2016. After applying exclusion criteria, 111 patients were identified for inclusion in this study. Patients were stratified by the timing of their intervention into immediate management (IM), within 24 hours and delayed management (DM), after 24 hours of their referral. Of the 111 poor-grade patients enrolled on this study, 53 (47.7%) received intervention within 24 hours (IM), and 58 (52.3%) received the intervention after 24 hours from the referral (DM). The mean time delay from referral to intervention was 14.9±5.8 hours in IM patients, compared to 79.6±106.1 hours in DM patients. The baseline characteristics of the IM and DM groups are shown in **Table 1**. Patient sex, GCS score, presence of hydrocephalus and pupillary deficit did not differ significantly between groups. However, there were significant differences in patient age (p=0.021), CCI score (p=0.037), GCS score (p=0.012) and intervention type received (p=0.045). The IM group had a significantly higher proportion of clipped patients than the DM group (17.0% vs 5.17%, p=0.045).

| Characteristic |  | IM (n=53) | DM (n=58) | P-value |
| --- | --- | --- | --- | --- |
| Male (%) |  | 17 (32.1) | 20 (34.5) | 0.788* |
| Age (±SD) | | 53.2 (11.9) | 58.4 (11.8) | 0.021** |
| CCI score [Median (IQR)] |  | 2 (1,3) | 3 (2,3.75) | 0.037*** |
| GCS score [Median (IQR)] |  | 9 (8,11) | 7 (5,10) | 0.012*** |
| Hydrocephalus (%) |  | 11 (20.8) | 20 (34.5) | 0.162* |
| Pupillary deficit (%) |  | 8 (15.1) | 15 (25.9) | 0.107* |
| Intervention received (%) | Clipping | 9 (17.0) | 3 (5.17) | 0.045* |
|  | Coiling | 44 (83.0) | 55 (94.8) |  |
| Time until management in hours (±SD) | | 14.9 (5.8) | 79.6 (106.1) | <0.001** |

\*Chi-squared test
\*\*T-test
\*\*\*Mann Whitney test
IM – Immediate management
DM – Delayed management
SD – Standard deviation
IQR – Interquartile range
CCI – Charlson Comorbidity Index
GCS – Glasgow Coma Scale

### Primary Outcome

At 12 months following discharge, there was no significant difference between the groups in the proportion of patients with favourable outcome (adjusted OR 0.96 [CI 0.17, 5.39; p=0.961]).

### Secondary Outcomes

At discharge, IM patients were significantly more likely to have favourable outcome than DM patients (adjusted OR 3.85 [CI 1.38, 10.73; p=0.010]). However, there was no difference between the groups in the likelihood of favourable outcome at three months (adjusted OR 1.04 [CI 0.22, 5.00; p=0.956]) and six months (adjusted OR 0.98 [CI 0.21, 4.58; p=0.982]).

Survival rate did not differ significantly between the two groups at 28 days, three months, six months and 12 months. The survival rate at 12 months in the IM and DM groups was 71.7% and 82.8%, respectively (p=0.163) (Table 3).

| Time | Odds Ratio<br>(95% CI) | P-value | Odds Ratio<sup>#</sup><br>(95% CI) | P-value | Number of patients<br>reported (%) |
| --- | --- | --- | --- | --- | --- |
| **Hospital discharge** |  |  |  |  | **87 (96.7)** |
| DM (reference) | 1 |  |  |  | 48 (98.0) |
| IM | 2.83 (1.17,<br>6.87) | 0.021 | 3.85 (1.38,<br>10.73) | 0.010 | 39 (95.1) |
| **3 months** |  |  |  |  | **54 (60.7)** |
| DM (reference) | 1 |  |  |  | 28 (58.3) |
| IM | 1.40 (0.38,<br>5.12) | 0.611 | 1.04 (0.22,<br>5.00) | 0.956 | 26 (63.4) |
| **6 months** |  |  |  |  | **54 (62.1)** |
| DM (reference) | 1 |  |  |  | 24 (50.0) |
| IM | 0.86 (0.24,<br>3.17) | 0.826 | 0.98 (0.21,<br>4.58) | 0.982 | 30 (76.9) |
| **12 months** |  |  |  |  | **55 (64.0)** |
| DM (reference) | 1 |  |  |  | 31 (64.6) |
| IM | 0.73 (0.18,<br>2.89) | 0.654 | 0.96 (0.17,<br>5.39) | 0.961 | 24 (63.2) |

Odds Ratio (OR): Modelling favourable outcome (unfavourable, favourable)
IM – Immediate management
DM – Delayed management
GOS – Glasgow Outcome Scale

- Adjusted Odds ratio (Age, Gender, CCI, GCS and Intervention received)
| **Time after haemorrhage** | **IM** | **DM** | **P-value** |
| --- | --- | --- | --- |
| **28 days** | 41 (77.4) | 49 (84.5) | 0.338 |
| **3 months** | 41 (77.4) | 48 (82.8) | 0.476 |
| **6 months** | 39 (73.6) | 48 (82.8) | 0.241 |
| **12 months** | 38 (71.7) | 48 (82.8) | 0.163 |

## DISCUSSION

Our study aimed to determine whether immediate management within 24 hours (IM) or delayed management after 24 hours (DM) was associated with better patient outcomes in pSAH. Our centre offers 24/7 coiling and clipping services, a practice which was in place throughout the study period. To our knowledge, we report on the largest cohort of pSAH patients to date investigating the effects of intervention time on long-term outcomes. The primary outcome was disability status of survivors (GOS 2-5), expressed as odds ratio of good outcome (GOS4-5). GOS 1 (dead) patients were analysed separately as a secondary outcome. We found that intervention within 24 hours was associated with comparable patient outcomes to intervention after 24 hours at 12 months. IM was associated with lower disability at discharge than DM, but this difference did not persist after three months. No significant differences were found in survival rate between IM patients and DM patients.

A recent observational study by Tykocki et al.^13^ in a cohort of 79 poor-grade patients found that IM is associated with better outcomes on discharge than DM. Our study adds to these results by reporting on a larger cohort and following up patient outcomes up to 12 months. Our study demonstrates that these observed improvements in outcomes are not preserved after three months.

Retrospective cohort studies are often at risk of confounding factors. However, our tertiary neurosciences database has a complete dataset of patient demographics, clinical condition, comorbidities, and relevant investigations for all patients. Hence, data on baseline characteristics were available for all patients included in this study, allowing us to statistically model the differences between the IM and DM groups at baseline and adjust the analysis of GOS scores. We found that IM was associated with lower age and CCI score than DM, which reflects current practice in our unit of prioritising earlier intervention to those perceived to have a better prognosis. This introduces a likelihood of a selection bias that would be expected to favour better outcomes in the IM cohort.

As this was a retrospective study, it was not possible to take measures to minimise the absence of patient’s disability data after discharge. As a result, there was a substantial loss of data during follow-up. While data on disability status were available for almost all patients who survived to discharge (87 out of 90, 96.7%), at three months, six months and 12 months, disability data were available in only 60.7%, 62.1% and 64.0% of alive patients, respectively. This may have introduced observation bias towards lower GOS scores for discharged patients, as patients with a higher disability often received more clinic appointments. Despite high rates of loss to follow-up, this study managed to report disability status for over 50 pSAH patients at each time point, up to 12 months following the discharge (Table 2).

The primary outcome was measured at discharge and fixed time intervals following discharge, rather than the ictus. This does not account for the patient’s length of stay, which may have differed between the IM and DM groups. Such a difference would mean the fixed time intervals following discharge would correspond to different lengths of time following SAH in the groups.

Our study provides supporting evidence that pSAH patients report promising rates of good functional recovery following discharge. This finding is in agreement with results from other recent observational studies on poor-grade patients, which showed a continuous improvement of neurocognitive function after discharge.^12-14^ This emerging evidence strongly supports the decision to intervene on pSAH and patients should no longer be rejected for definitive treatment based on a poor clinical grade at presentation.^11,16,17^ In our unit, in 2011, only 27.0% of pSAH patients received definitive treatment to secure their aneurysm; by 2016, this proportion had doubled to 64.3%.

Early intervention has become the routine in managing patients with aneurysmal SAH.^20-23^ While this reduces the risk of re-bleed, pragmatically, immediate management does not allow effective planning of the intervention. Our results indicate that immediate intervention within 24 hours is not associated with significant improvements in morbidity or mortality at 12 months in pSAH patients compared to delayed intervention after 24 hours. This is despite the likelihood of a selection bias favouring earlier intervention in patients perceived to have a better prognosis. Delaying intervention allows for patient management to be appropriately discussed in a multidisciplinary setting with involvement from neuro-critical care physicians, neuro-anaesthetists, interventional neuro-radiologists and neurosurgeons. The procedure can also be scheduled for a time as soon as possible that maximises the availability of resources, ensuring optimal patient care. Although intervention within 24 hours in pSAH has been shown to produce good outcomes, it is associated with very high perioperative mortality. Gupta et al.^14^ reported a mortality rate of 50.9% among poor-grade patients receiving intervention within 24 hours, while Laidlaw et al.^12^ found that nonselective urgent surgery within 12 hours was associated with a 45% mortality rate. The findings of our study indicate that delayed intervention may be an option in pSAH management without an increase in morbidity or mortality. Delaying intervention may give the critical care team time to stabilise the patient, thereby reducing overall surgical risk.

The observational nature of this study prevents definitive conclusions from being drawn about the optimal timing of intervention in pSAH. In the UK, the results of our research should have been taken together with results of the TOPSAT2 study^5^, which has now stopped due to the slow recruitment. Further research to clarify the optimal time window to intervene after pSAH is required.

## CONCLUSIONS

IM and DM after pSAH are associated with similar morbidity and mortality. Given that IM and DM have similar outcomes at 12 months, it may be appropriate to delay intervention if clinically indicated to stabilise the patient and adequately plan the procedure.

## Data Availability

None required

## Acknowledgements

The manuscript complies with all instructions to authors. All authors have read and approved the submitted manuscript; the manuscript has not been submitted elsewhere, nor published elsewhere in whole or in part.

---

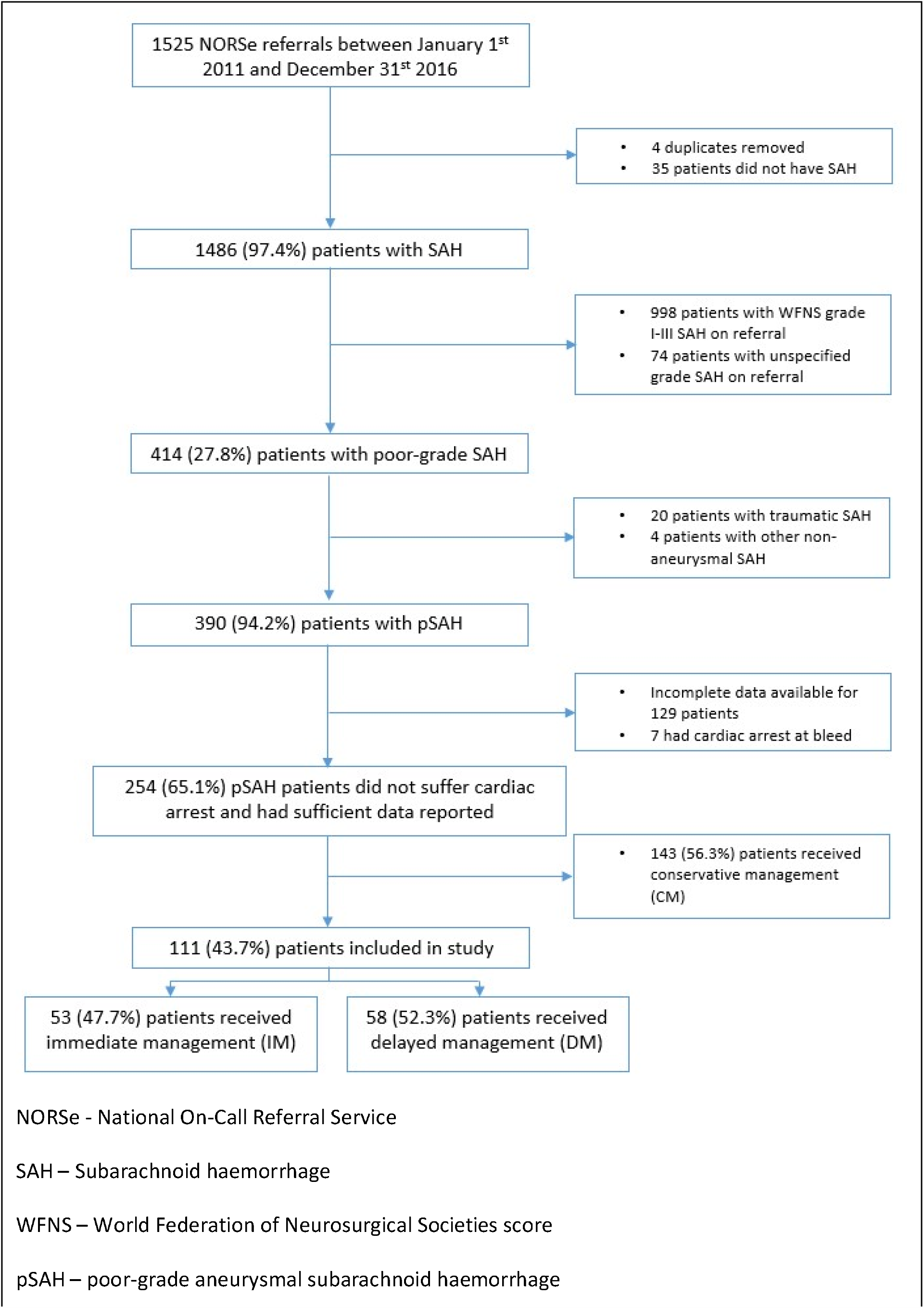

